# Predictive Modeling of Healthcare Workers’ Priorities of WHO 2030 Key Activities for Snakebite Prevention and Control in Ghana

**DOI:** 10.64898/2026.01.13.26344079

**Authors:** Eric Nyarko, Iddrisu Abugbil Atubiga, Fafa Shalom Nkunim Tchorly, Nicholas Amani Hamman, Aashna Uppal, Nuhu Mohammed, Eduardo Alberto Fernandez, Isaac Baidoo

## Abstract

Snakebite is a neglected public health problem that results in significant morbidity and mortality, necessitating the World Health Organization (WHO) to develop a snakebite roadmap that aimed to halve the burden of snakebite envenoming (SBE) by 2030. This study predicted healthcare workers’ priorities regarding the WHO’s 2030 snakebite strategic key activities for preventing and controlling SBE. This cross-sectional study was conducted in the Kwahu Afram Plains North and South districts of Ghana from August to December 2024 and involved 137 healthcare workers, including physici n assistants, clinical officers, medical doctors, certificate/enrolled/general nurses, pharmacists, dispensing technicians, and community health nurses, using a multistage sampling technique. Maximum difference choice experiments and five machine learning models were used to analyze the data. Healthcare workers prioritized the activity “Making safe, effective antivenoms available, accessible, and affordable to all” as the most crucial key activity, with a utility estimate (UE) of 0.8256 (95% confidence interval (CI): 0.7300 to 0.9213), followed by “Effective first aid care and ambulance transport” (UE = 0.4348, 95% CI: 0.3266, 0.5430), “Coordinated data management and analysis” (UE = 0.3744, 95% CI: 0.2353, 0.5134), and “Promoting advocacy, effective communication, and productive engagement” (UE = 0.3630, 95% CI: 0.2528, 0.4732). The use of choice experiments and ML models has provided insights into healthcare workers’ priorities concerning the WHO’s 2030 key activities for snakebite prevention and control. This innovative approach offers a nuanced understanding of local perspectives on WHO’s key activities, which is essential for combating the burden of SBE in Ghana.

## 1. Introduction

Globally, approximately 5.8 billion people are at risk of venomous snakebites. Every day, about 7,400 individuals are bitten by snakes [1], resulting in an annual death toll of between 81,000 and 138,000. Additionally, around 400,000 victims suffer from permanent physical or psychological disabilities, which can include blindness, tissue loss, amputations, and post-traumatic stress disorder [2]. In northern Ghana, the reported incidence of snakebites is 56.4 cases per 100,000 inhabitants, with a mortality rate of 1.35 per 100,000 [3]. In the Volta region, data from health facilities indicate an incidence rate of 15.8 snakebite cases per 100,000 population and a case fatality rate of 0.4% [4].

As efforts to achieve the objectives of UHC2030 (https://www.uhc2030.org/) progress, immediate action is essential to alleviate the suffering of the world’s most disadvantaged communities [5]. Recognizing the critical public health issue of snakebite, the World Health Organization (WHO) classified snakebite envenoming (SBE) as a Category A neglected tropical disease (NTD) in 2017 and a resolution at the 2018 World Health Assembly [6], leading to the development of a global strategy with four key objectives. Each objective includes six key activities aimed at halving the number of deaths and disabilities caused by SBE by 2030 [5]. Achieving this goal relies heavily on high-quality research evidence to guide effective policies and interventions [7, 8]. This need extends to designing and implementing locally relevant plans to combat SBE [1]. Therefore, rigorous and innovative research is essential to inform the development of such localized plans or policies. These initiatives will help identify regional priorities that shape local SBE research agendas and develop interventions that address community needs. The cutting-edge technology of artificial intelligence (AI) holds significant potential in this area of tropical disease [9], offering hope and optimism for the future of SBE management.

Numerous studies have utilized AI models to help identify and classify snakes and predict snakebites [10, 11, 12, 13, 14] without exploring statistical experiment designs. Combining AI or machine learning (ML) models with statistical experimental design can yield valuable insights to enhance data-driven decision making [15]. This study establishes the foundation for the exploration of maximum difference choice experiments techniques, employing combinatorial methods such as fractional factorial designs, incomplete block designs, and Hadamard matrices, in conjunction with AI/ML applications in SBE. A previous study in Ghana used a qualitative approach to rank the six key activities outlined in each of WHO’s SBE four strategic objectives [7]. This study conducted in Ghana’s Kwahu Afram Plains North and South districts on account of reported snakebite cases and related fatalities [16] supports the existing literature by providing quantitative, evidence-based insights based on the WHO’S key activities. This study aimed to predict healthcare workers’ priorities regarding the WHO’s 2030 snakebite strategic key activities for preventing and controlling SBE. Our prior studies quantified healthcare workers’ priorities regarding the WHO’s four strategic objectives [17]. In the current study, our goal is to provide a robust platform through rigorous research evidence based on the WHO’S key activities that can guide the development of SBE research agendas and relevant interventions or policies that prioritize local needs. By integrating this information into the health system, there is potential to significantly improve health outcomes and the quality of life for snakebite patients in Ghana and other low- and middle-income countries (LMICs).

## 2. Methods

### 2.1. Study design

This cross-sectional study was part of a large survey carried out from August to December 2024 in the Kwahu Afram Plains North and South districts in Ghana’s Eastern Region, where snake bites and related deaths have been reported [16]. We employed a multi-stage sampling technique and interviewer-administered questionnaires to identify healthcare workers involved in the treatment and management of snakebite patients in these districts and to assess their priorities regarding the WHO’s strategic objectives’ key activities for controlling and preventing SBE.

### 2.2. Designed maximum difference statistical choice experiment

This study employed a maximum difference choice experiment approach, an advanced method for assessing preferences for various goods and services [18, 19]. Experimental design is crucial for generating choice sets used in maximum difference studies. A well-structured statistical experiment design helps identify the specific choices for the task, which are defined by a number of attributes and their corresponding levels. This method allows for effective manipulation of attributes and levels, enabling thorough testing of research objectives and instilling confidence in the study’s methodology. There were 1,296 experimental conditions based on the WHO’s SBE four strategic objectives and their associated six key activities. However, creating choice sets of size three from these experimental conditions would have resulted in an impractically large number of scenarios. To ensure the study’s efficiency and to prevent information overload for the respondents, we utilized combinatorial methods, including fractional factorial designs, incomplete block designs, and Hadamard matrices, to condense the scenarios into 12 choice sets of size three required for a best-worst scaling task. We assume that these choice sets have equal chance of being selected. In these effects-type coded choice sets, the frequency of the first attribute’s six levels (L1 to L6) appeared as follows: L1 = 6, L2 = 6, L3 = 5, L4 = 8, L5 = 6, L6 = 5. The frequency of the levels of the second attribute appeared as: L1 = 5, L2 = 7, L3 = 7, L4 = 6, L5 = 7, L6 = 4. The frequency of the levels of the third attribute appeared as: L1 = 7, L2 = 8, L3 = 4, L4 = 6, L5 = 5, L6 = 6, while the frequency of the levels of the fourth attribute appeared as: L1 = 8, L2 = 9, L3 = 5, L4 = 6, L5 = 5, L6 = 3. Fig. 1 shows a sample choice set.

**Fig. 1.**
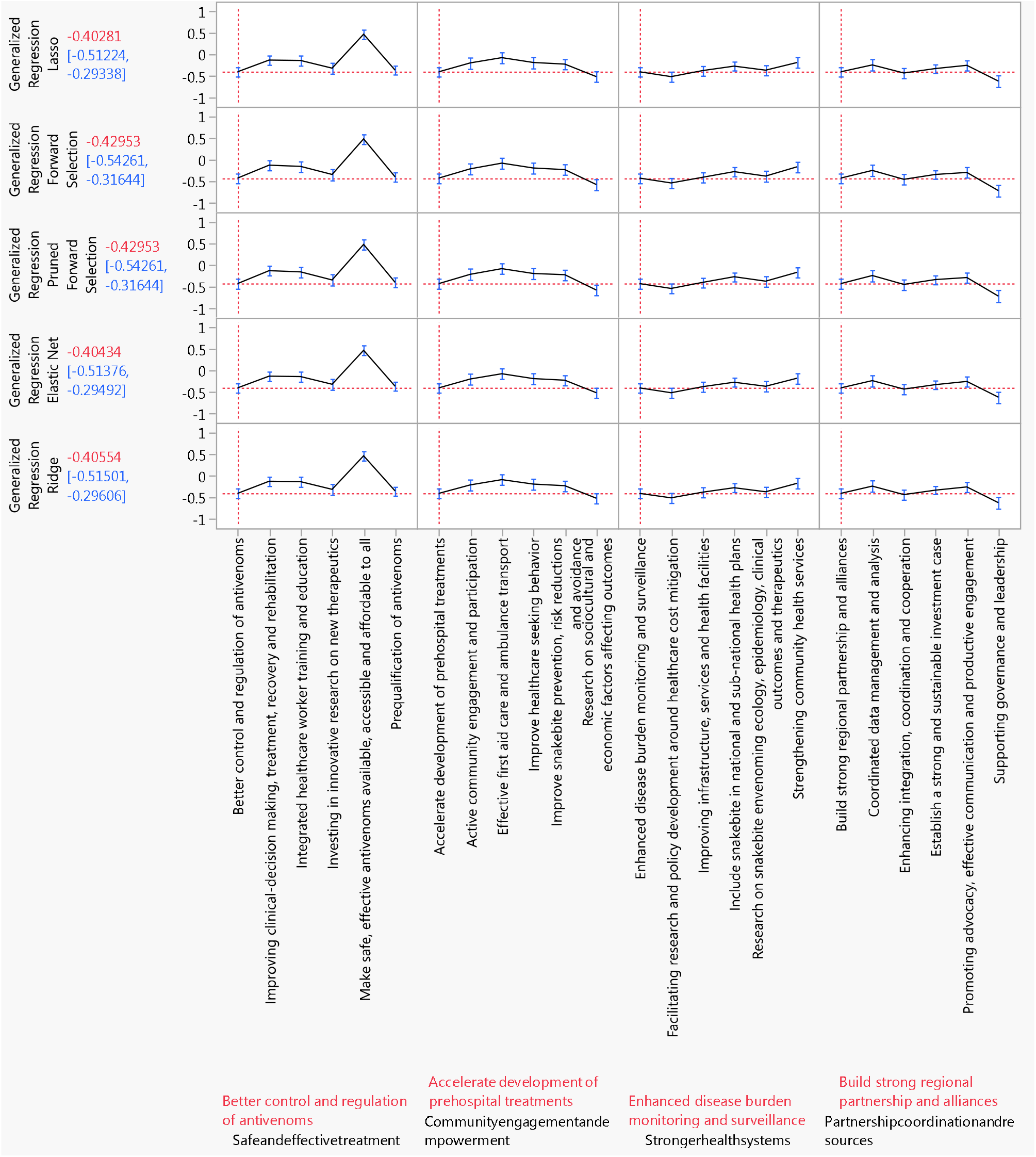
Prediction profiler for WHO SBE strategic objectives’ key activities.

### 2.3. Sample size and data collection

In this study, the sample size was calculated using the available scenarios (l=12) and the formula *N=n/l*, for any given sample *n* > *r*^*2*^*s/va*^*2*^ [20]. In this formula, r represents the inverse cumulative distribution function of a standard normal distribution, v is the true choice proportion of the study population based on the assumption of equal choice probabilities, and a is the allowable deviation as a percentage between the estimated proportion v” and v. Additionally, s = 1 - v, and a signifies a 95% confidence level. This assessment indicated that a minimum of 128 respondents were needed for our study. We ultimately decided to collect data from 137 respondents. We tested the survey tool in a pilot study with twenty healthcare professionals to ensure it was clear. During the survey, respondents were presented with 12 choice sets through an interviewer-administered questionnaire. They were asked to select their most and least prioritized needs concerning the key activities of the WHO’s SBE strategic objectives. Participants recorded their responses using paper and pencil. Two enumerators entered the data using Microsoft Excel before it was imported into JMP Pro (Version 17.0) for formal analysis.

### 2.4. Data preparation for model building

Data collected during our field survey indicated no issues with outliers or missing values because enumerators encouraged respondents to complete any unfinished sections of the survey tools. We employed the hold-back validation method to enhance the model’s generalization ability and reduce the risk of overfitting. This method involves splitting the data into training and validation sets, with 70% allocated for training and 30% for validation. It is important to note that the predictor variables, which are associated with the key activities of the WHO SBE strategic objectives, were treated as generic attributes, while the response variable was treated as continuous.

### 2.5. Data management and analysis

We analyzed the dataset using five ML models: Ridge Regression [21], Elastic Net [22], least absolute shrinkage and selection operator (LASSO) [23], a Generalized Regression Model with Pruned Forward Selection, and Forward Selection [24]. To compare the performance of these models, we utilized several key metrics, including the corrected Akaike Information Criterion (AICc), the Bayesian Information Criterion (BIC), the Root Average Squared Error (RASE), negative log-likelihood, and the elapsed time to fit each model. All predictor variables were deemed statistically significant if the p-value was less than or equal to 0.05 (with varying degrees of significance if the p-value was less than 0.001 or 0.01), or if the 95% confidence intervals (CIs) did not include zero. All statistical analyses were conducted using JMP Pro (Version 17.0).

## 3. Results

### 3.1. Participant characteristics

One hundred thirty-seven individuals participated in this study, representing a diverse range of healthcare professionals as detailed in a previous study. Among them, 96 (70.0%) were Certified, Enrolled, or General Nurses, reflecting strong nursing representation. The study also included a variety of other healthcare roles, with 16 participants (11.7%) being Physician Assistants, Clinical Officers, and Medical Doctors, 12 (8.8%) being Pharmacists or Dispensing Technicians, and 13 (9.5%) being Community Health Nurses. This diversity in roles informs the breadth of the study’s scope and the range of perspectives considered. Most participants were experienced, with 98 individuals (71.5%) aged 31 or older. Gender representation included 83 females (60.6%), and 60 participants (43.8%) reported having at least six years of work experience, highlighting a wealth of practical knowledge in the cohort.

### 3.2. Model fit and overall performance

Table 2 compares the ML models used in this study based on various evaluation metrics. The LASSO model achieved the lowest AICc of 3426.8732 and BIC of 3537.5352. Despite requiring the longest time of 4945 milliseconds for fitting, it demonstrated the best predictive performance with an RASE of 0.7589095 and a negative log-likelihood of 1692.1198. This performance highlights the trade-off between computational efficiency, goodness of fit, and predictive accuracy, a crucial factor in the model selection process. The Elastic Net regression model followed closely, with an AICc of 3426.8841, a BIC of 3537.5461, a RASE of 0.7589097, a negative log-likelihood of 1692.1252, and a better fitting time of 4022 milliseconds compared to the LASSO model. The Generalized Regression (Forward Selection) model was the most efficient, taking a fitting time of just 224 milliseconds, while the Pruned Forward Selection model took 601 milliseconds. Still, they had the highest AICc of 3431.9669 and a RASE of 0.7596380. In conclusion, the LASSO model is the best choice, achieving the lowest metrics for RASE, AICc, and BIC, which makes it ideal for applications prioritizing predictive accuracy.

**Table 1.**
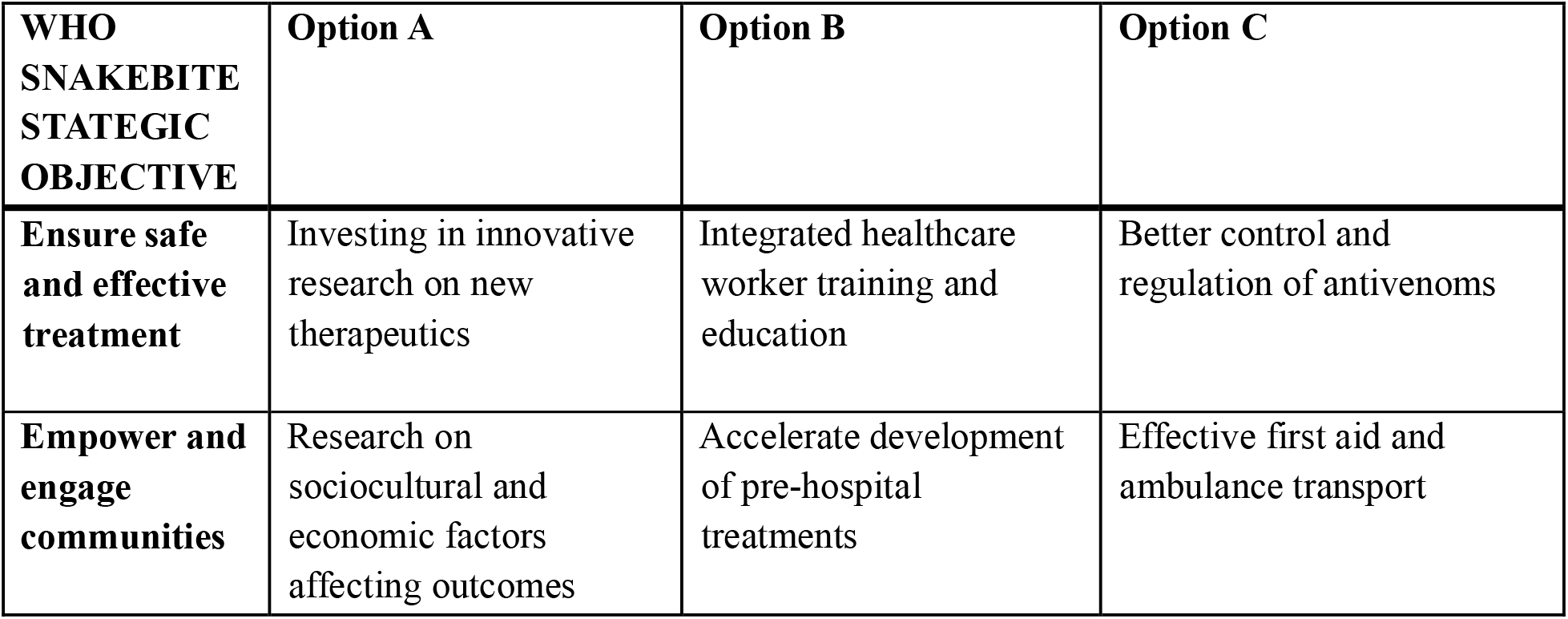
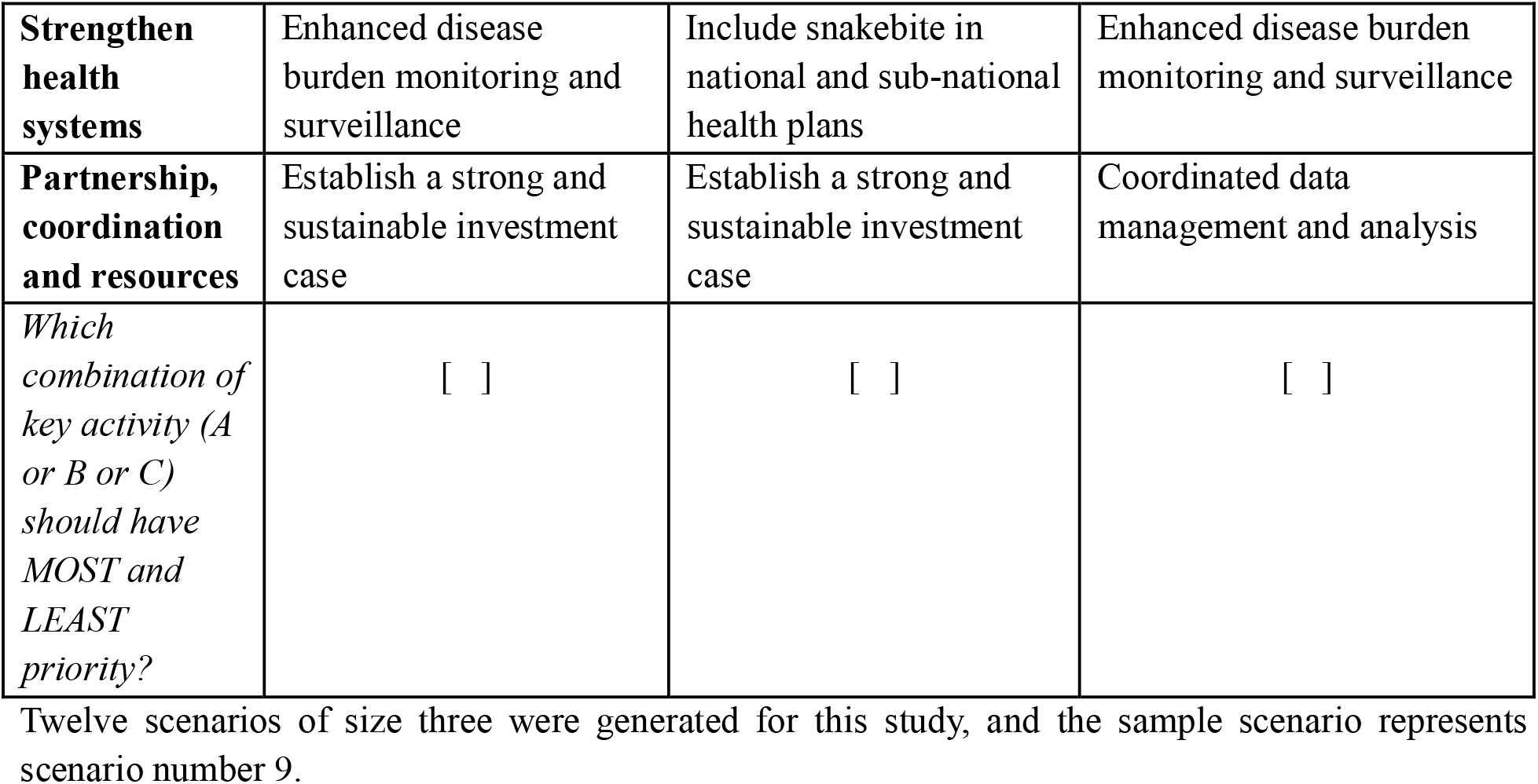
Example of one of the scenarios as it appeared in the survey.

**Table 2.** Model performance evaluations.

| **Method** | **AICc** | **BIC** | **RASE** | **Elapsed time**<br>**(milliseconds**<br>**)** | **Negative**<br>**log-**<br>**likelihood** |
| --- | --- | --- | --- | --- | --- |
| LASSO | 3426.8732 | 3537.5352 | 0.7589095 | 4945 | 1692.1198 |
| Generalized regression<br>model (Forward selection) | 3431.9669 | 3547.8678 | 0.7596380 | 224 | 1693.6361 |
| Generalized regression<br>model (Pruned forward<br>selection) | 3431.9669 | 3547.8678 | 0.7596380 | 601 | 1693.6361 |
| Elastic Net | 3426.8841 | 3537.5461 | 0.7589097 | 4022 | 1692.1252 |
| Ridge | 3427.1392 | 3537.8012 | 0.7589715 | 3208 | 1692.2527 |

AICc: corrected Akaike Information Criterion; BIC: Bayesian Information Criterion; RASE: Root Average Squared Error; LASSO: Least Absolute Shrinkage and Selection Operator.

### 3.3. Prediction profilers for each type of model

Fig. 1 visually represents the utility profilers for each type of ML model. These profilers are essential in demonstrating the importance assigned to the key activities of the WHO’s strategic objectives for preventing and controlling SBE. The vertical red line in the utility profiler indicates the current value of these strategic objectives’ key activities, while the horizontal red lines show the current predicted values with their respective CIs of each response variable based on those key activities. The ML models consistently identified the key activities as significant strategies for preventing and controlling SBE, with a few exceptions. Specifically, the key activities “Better control and regulation of antivenoms,” “Investing in innovative research on new therapeutics,” and “Include snakebite in national and sub-national health plans” were not identified as significant strategies. However, it is noteworthy that “Include snakebite in national and sub-national health plans” was recognized as a significant strategy by the Generalized Regression model (specifically using Pruned Forward Selection and Forward Selection methods).

### 3.4. LASSO Regression model

We presented the results from the LASSO regression model because it exhibited the best overall performance among all the candidate models. Utility estimates based on twenty key activities alongside the WHO SBE four strategic objectives are presented in Table 3. It is worth noting that the key activities “Prequalification of antivenoms,” “Research on sociocultural and economic factors affecting outcomes,” “Strengthening community health services,” and “Supporting governance and leadership” were treated as reference levels. Among the WHO SBE strategic objectives’ key activities, the model identifies “Make safe, effective antivenoms available, accessible, and affordable to all” as the most significant key activity (utility estimate (UE) = 0.8256, 95% CI: 0.7300, 0.9213) relative to the reference levels. Other WHO SBE key activities of high significance include “Effective first aid care and ambulance transport” (UE = 0.4348, 95% CI: 0.3266, 0.5430), followed by “Coordinated data management and analysis” (UE = 0.3744, 95% CI: 0.2353, 0.5134), “Promoting advocacy, effective communication, and productive engagement” (UE = 0.3630, 95% CI: 0.2528, 0.4732), “Improve healthcare-seeking behavior” (UE = 0.3182, 95% CI: 0.2123, 0.4242), “Active community engagement and participation” (UE = 0.3087, 95% CI: 0.2123, 0.4050), “Establish a strong and sustainable investment case” (UE = 0.2848, 95% CI: 0.1818, 0.3878), “Improve snakebite prevention, risk reduction, and avoidance” (UE = 0.2842, 95% CI: 0.1866, 0.3818), “Improving clinical decision-making, treatment, recovery, and rehabilitation” (UE = 0.2302, 95% CI: 0.1354, 0.3250), “Integrated healthcare worker training and education” (UE = 0.2190, 95% CI: 0.1239, 0.3140), “Build strong regional partnerships and alliances” (UE = 0.2129, 95% CI: 0.1094, 0.3164), “Enhancing integration, coordination, and cooperation” (UE = 0.1856, 95% CI: 0.0811, 0.2902), and “Accelerate the development of prehospital treatments” (UE = 0.1095, 95% CI: 0.0304, 0.1886).

**Table 3.** LASSO regression model results for WHO SBE strategic objectives’ key activities.

| Attribute | Utility estimate | SE | Wald ChiSquare | Prob > ChiSquare | Lower 95% | Upper 95% |
| --- | --- | --- | --- | --- | --- | --- |
| **Ensure safe and effective treatment:** |  |  |  |  |  |  |
| Better control and regulation of antivenoms | -0.0388 | 0.0421 | 0.8493 | 0.3567 | -0.1215 | 0.0438 |
| Improving clinical-decision making, treatment, recovery and rehabilitation | 0.2302 | 0.0483 | 22.6625 | <.0001* | 0.1354 | 0.3250 |
| Integrated healthcare worker training and education | 0.2190 | 0.0485 | 20.3830 | <.0001* | 0.1239 | 0.3140 |
| Investing in innovative research on new therapeutics | 0.0400 | 0.0568 | 0.4967 | 0.4809 | -0.0713 | 0.1515 |
| Make safe, effective antivenoms | 0.8256 | 0.0487 | 286.3552 | <.0001* | 0.7300 | 0.9213 |
| available, accessible and affordable to all |  |  |  |  |  |  |
| **Empower and engage communities:** |  |  |  |  |  |  |
| Accelerate development of prehospital treatments | 0.1095 | 0.0403 | 7.3625 | 0.0067* | 0.0304 | 0.1886 |
| Active community engagement and participation | 0.3087 | 0.0491 | 39.4144 | <.0001* | 0.2123 | 0.4050 |
| Effective first aid care and ambulance transport | 0.4348 | 0.0551 | 62.0828 | <.0001* | 0.3266 | 0.5430 |
| Improve healthcare seeking behavior | 0.3182 | 0.0540 | 34.6708 | <.0001* | 0.2123 | 0.4242 |
| Improve snakebite prevention, risk reductions and avoidance | 0.2842 | 0.0498 | 32.5635 | <.0001* | 0.1866 | 0.3818 |
| **Strengthen health systems:** |  |  |  |  |  |  |
| Enhanced disease burden monitoring and surveillance | -0.2150 | 0.0547 | 15.4199 | <.0001* | - 0.3223 | - 0.1077 |
| Facilitating research and policy development around healthcare cost mitigation | -0.3259 | 0.0559 | 33.9305 | <.0001* | - 0.4355 | - 0.2162 |
| Improving infrastructure, services and health facilities | -0.1921 | 0.0491 | 15.2960 | <.0001* | - 0.2884 | - 0.0958 |
| Include snakebite in national and sub-national health plans | -0.0835 | 0.0546 | 2.3407 | 0.1260 | - 0.1906 | 0.0234 |
| Research on snakebite envenoming ecology, epidemiology, clinical outcomes and therapeutics | -0.1766 | 0.0640 | 7.6010 | 0.0058* | - 0.3022 | - 0.0510 |
| **Partnership, coordination and resources:** |  |  |  |  |  |  |
| Build strong regional partnership and alliances | 0.2129 | 0.0528 | 16.2518 | <.0001* | 0.1094 | 0.3164 |
| Coordinated data management and analysis | 0.3744 | 0.0709 | 27.8461 | <.0001* | 0.2353 | 0.5134 |
| Enhancing integration, coordination and cooperation | 0.1856 | 0.0533 | 12.1224 | 0.0005* | 0.0811 | 0.2902 |
| Establish a strong and sustainable | 0.2848 | 0.0525 | 29.3673 | <.0001* | 0.1818 | 0.3878 |
| investment case |  |  |  |  |  |  |
| Promoting advocacy, effective communication and productive engagement | 0.3630 | 0.0562 | 41.7115 | <.0001* | 0.2528 | 0.4732 |
| **Model Fit Statistics** |  |  |  |  |  |  |
| AICc | 3426.8732 |  |  |  |  |  |
| BIC | 3537.5352 |  |  |  |  |  |
| RASE | 0.7589095 |  |  |  |  |  |
| Negative Log-Likelihood | 1692.1198 |  |  |  |  |  |

AICc: corrected Akaike Information Criterion ; BIC: Bayesian Information Criterion; RASE: Root Average Squared Error; SE: Squared Error

While statistically significant, participants tend to trade off certain key activities. These include “Facilitating research and policy development around healthcare cost mitigation” (UE = −0.3259, 95% CI: −0.4355, −0.2162), “Enhanced disease burden monitoring and surveillance” (UE = −0.2150, 95% CI: −0.3223, −0.1077), “Improving infrastructure, services, and health facilities” (UE = −0.1921, 95% CI: −0.2884, −0.0958), and “Research on snakebite envenoming ecology, epidemiology, clinical outcomes, and therapeutics” (UE = −0.1766, 95% CI: −0.3022, −0.0510).

## 4. Discussion

This study employed innovative and advanced research methods, including the maximum difference statistical experiment design and ML models to rigorously predict healthcare workers’ priorities regarding key activities under the WHO strategic objectives for controlling and preventing SBE. A key finding of our study was that healthcare workers prioritized “Make safe, effective antivenoms available, accessible, and affordable to all” as the most significant key activity. Other significant priorities included “Effective first aid care and ambulance transport”, “Coordinated data management and analysis”, “Promoting advocacy, effective communication, and productive engagement”, “Improving healthcare-seeking behavior”, “Active community engagement and participation”, “Establishing a strong and sustainable investment case”, “Enhancing snakebite prevention, risk reduction, and avoidance”, “Improving clinical decision-making, treatment, recovery, and rehabilitation”, “Integrating healthcare worker training and education”, “Building strong regional partnerships and alliances”, “Enhancing integration, coordination, and cooperation”, and “Accelerating the development of prehospital treatments”.

Our findings align with a previous study in Ghana [7], reinforcing the importance of the key activity “Make safe, effective antivenoms available, accessible, and affordable to all.” This consistency provides a solid foundation for formulating local and regional policies or interventions and setting research agendas. Prior studies have advocated for solutions that address practical problems, yield innovative solutions, enhance the potency, specificity, and safety of current antivenoms, and lead to effective treatment, faster recovery, fewer deaths, and lower long-term disability [25, 26]. Direct action—including global risk-benefit assessments of antivenom products to determine their suitability based on risks versus benefits and issues in antivenom production—is necessary to reshape regional antivenom markets [26]. The high cost of antivenoms, the lack of access to effective antivenoms in remote areas, the increased use of unorthodox and harmful practices, and the unavailability of effective antivenoms for local species have been cited as challenges that limit the accessibility and utilization of antivenoms for treating SBE in Ghana [27]. However, implementing WHO-recommended products through a revolving supply facility and establishing an antivenom stockpile program is commendable and could help restore confidence in using antivenoms [26].

Under the second objective, “Empower and engage communities”, the activity “Active community engagement and participation” was cited in a previous study as a top priority at the community level in Ghana [7]. This finding contrasts with our results, where “Effective first aid care and ambulance transport” was prioritized under the second objective. It has been noted that the time it takes for a snakebite victim to reach a medical facility with regional antivenom can range from hours to days, with long travel times leading to increased mortality and morbidity related to SBE [28, 29]. There is an urgent need to equip communities with the best first aid practices and essential actions that improve victims’ health and survival immediately after a snakebite while organizing transportation to health facilities as quickly as possible. An excellent resource is the essential first-aid procedures outlined in the WHO Guidelines for Preventing and Clinical Managing Snakebites in Africa [30].

In a previous study [7], the activity “Promoting advocacy, effective communication, and productive engagement” was ranked as a top priority under the strategic objective of “Partnership, coordination, and resources.” This result contrasts with our present study, where “Coordinated data management and analysis” emerged as the top priority. The significance of this finding is emphasized by previous research that highlights the importance of having access to accurate information, research data, and surveillance results. These elements are fundamental to health planning, monitoring, and assessment and are crucial for a strong health system and for eliminating NTDs [31, 32]. Prior studies have noted the relative lack of high-quality epidemiological surveillance data and the impact this has on accurately reporting the burden of disease [2, 33]. Therefore, it is imperative to enhance the surveillance of SBE in Ghana and other SSA countries. An excellent initiative is including SBE data in the Global Health Observatory (www.who.int/gho/), which will improve data quality and comparability. This includes standardizing clinical criteria adapted to specific regional needs and developing minimum data set definitions for community-acquired and hospital-acquired data to inform decision-making [5, 26].

In efforts to gather the evidence needed to support the WHO’s strategies for achieving a 50% reduction in global SBE mortality and disability, various activities have been prioritized. These include “Promoting advocacy, effective communication, and productive engagement,” “Improving healthcare-seeking behavior,” “Active community engagement and participation,” “Establishing a strong and sustainable investment case,” “Enhancing snakebite prevention, risk reduction, and avoidance,” and “Improving clinical decision-making, treatment, recovery, and rehabilitation.” Other priorities involve “Integrated healthcare worker training and education,” “Building strong regional partnerships and alliances,” “Enhancing integration, coordination, and cooperation,” and “Accelerating the development of prehospital treatments.” As we push forward with the objectives outlined in UHC2030 (https://www.uhc2030.org/), it is crucial to take immediate action to address these local priority needs through comprehensive programs. This will be fundamental in effectively reducing the global burden of SBE by 2030 and providing many of the world’s poorest and most vulnerable communities with the opportunity to lead healthy and productive lives [5, 26] Previous studies have also called for multidisciplinary research initiatives in the region [8] and established guidelines for training healthcare workers in the management of SBE [30].

A striking finding was that while participants acknowledged the significance of all key activities under the strategic objective “Strengthen health systems,” there was a tendency to trade off among them. These activities include “Facilitating research and policy development around healthcare cost mitigation,” “Enhanced disease burden monitoring and surveillance,” “Improving infrastructure, services, and health facilities,” and “Research on the ecology, epidemiology, clinical outcomes, and therapeutics of snakebite envenoming.” Managing snakebite envenoming is an acute emergency requiring responsive health systems to ensure timely service delivery [1]. Strengthening healthcare capacity and performance to manage SBE and other diseases at both community and national levels is vital and integral to achieving the goals outlined in UHC2030 [26, 34].

It is important to acknowledge the study’s limitations. The study was confined to a limited geographical area, primarily reflects the perspectives of healthcare workers, and did not explore interaction effects or subgroup analysis by cadre. Future comprehensive population-based surveys should incorporate the views of community members and other key stakeholders in healthcare delivery, health policy, research, and decision-making related to NTDs. There may be an issue of cognitive burden regarding the best–worst task.

In conclusion, this study offers valuable quantitative insights into healthcare workers’ priorities regarding the WHO’s 2030 key activities for snakebite prevention and control. This innovative approach provides a nuanced understanding of local perspectives on these activities, which is crucial for addressing the burden of SBE in Ghana.

## Consent for publication

Not applicable

## Data availability statement

Data will be made available on request.

## Clinical trial number

Not applicable.

## Ethical considerations

The study received ethical approval from the Ghana Health Service Ethics Review Committee (GHS-ERC073/04/24) and adhered to all ethical guidelines and regulations. After explaining the purpose of the study to all participants, written informed consent was obtained. Participants were informed that their participation was voluntary and that they had the option to choose whether to participate in the study or not.

## Acknowledgments

We would like to thank all the health workers who volunteered and shared their knowledge and experiences. We are grateful to Prof. José María Gutiérrez for providing comments on the manuscript.

